# HLA-C^*^ 04:01 is a Genetic Risk Allele for Severe Course of COVID-19

**DOI:** 10.1101/2020.12.21.20248121

**Authors:** Weiner January, Suwalski Phillip, Holtgrewe Manuel, Thibeault Charlotte, Melina Müller, Patriki Dimitri, Quedenau Claudia, Ulrike Krüger, T. Helbig Elisa, Lippert Lena, Stubbemann Paula, Real Luis Miguel, Sanchez Juan Macias, A. Pineda Juan, Fernandez-Fuertes Marta, Wang Xiaomin, Karadeniz Zehra, Saccomanno Jacopo, Doehn Jan-Moritz, Hübner Ralf-Harto, Hinzmann Bernd, Salvo Mauricio, Blueher Anja, Siemann Sandra, Jurisic Stjepan, Beer Hansjuerg, Rutishauser Jonas, Wiggli Benedikt, Schmid Hansruedi, Danninger Kathrin, Binder Ronald, M. Victor Corman, Mühlemann Barbara, Braun Alice, Ripke Stephan, C. Jones Terry, Suttorp Norbert, Witzenrath Martin, Hippenstiel Stefan, Zemojtel Tomasz, Skurk Carsten, Poller Wolfgang, Borodina Tatiana, Pa-COVID Study Group, E. Sander Leif, Beule Dieter, Landmesser Ulf, Guettouche Toumy, Kurth Florian, Heidecker Bettina

**Author notes:** **Corresponding author:** Bettina Heidecker, MD, Charité Universitaetsmedizin Berlin, Campus Benjamin Franklin, Department of Cardiology, Hindenburgdamm 30, 12203 Berlin, Germany. **Pa-COVID-19 Study Group:** Mirja Mittermaier;Tilman Lingscheid; Pinkus Tober-Lau; Lil Meyer-Arndt; Sascha S.Haenel; Laure Bosquillon de Jarcy; Moritz Pfeiffer; Miriam S. Stegemann; Robert Roehle; Janine Wiebach; Thomas Zoller; Holger Müller-Redetzky; Alexander Uhrig; Felix Balzer; Christof von Kalle; Sascha Treskatsch; Stefan Angermair; Julia Heeschen; Linda Jürgens; Malte Kleinschmidt; Sophy Denker; Christoph Ruwwe-Glösenkamp; Bettina Temmesfeld-Wollbrück; Katrin M. Heim; Dirk Schürmann; Andreas Hocke; Bastian Opitz; Belén Millet Pascual-Leone; Rosa C. Schuhmacher; Nadine Olk; David Hillus; Felix Machleidt; Sebastian Albus; Felix Bremer; Carmen Garcia; Philipp Knape; Philipp M; Krause, Liron Lechtenberg; Yaosi Li; Panagiotis Pergantis; Teresa Ritter; Berna Yedikat; Christian Zobel; Friederike L. Hefele; Ute Kellermann; Mariana Schürmann; Lisa-Marie Wackernagel; Anne Wetzel; Daniel Grund; Jens K. Haumesser; Johannes Hodes; Johannes Rein; Peter Radünzel; Astrid Breitbart; Sergej Münzenberg; Dominik Soll; Tamar Zhamurashvili; Florian Alius; Tim Andermann; Thomas Cronen; Simon Fraumann; Nikolaj Frost; Dominik Geus; Gisele J. Godzick- Njomgang; Anne Herholz; Vera Hermanns; Moritz Hilbrandt; Till Jacobi; Ye-Ji Kim; Elena Madlung; Luise Martin; Nikolai Menner; Agata Mikolajewska; Luisa Mrziglod; Nadine Muller; Michaela Niebank; Eva Pappe; Frieder Pfäfflin; Lennart Pfannkuch; Matthias Raspe; Nicola Reck; Anne Ritter; Laura K. Schmalbrock; Fridolin Steinbeis; Christoph Tabeling; Markus Vogtmann; Susanne Weber; Markus Brack; Matthias Felten; Sein Schmidt; Maria Rönnefarth; Georg Schwanitz; Alexander Krannich; Saskia Zvorc; Uwe D. Behrens; Lucie Kretzler; Linna Li; Isabelle Wirsching; Chantip Dang-Heine; Michael Hummel; Dana Briesemeister; Denise Treue, Martin Möckel; Samuel Knauß; Matthias Endres; Claudia Spies; Steffen Weber; Carstens, Jan M. Kruse; Daniel Zickler; Andreas Edel; Britta Stier; Philipp Enghard; Roland Körner; Kai-Uwe Eckardt; Lucas Elbert; Christopher Neumann; Marius A. Eckart; Thuy N. Pham; Solveig Schönberger; Alexander Wree; Frank Tacke; Josef Mang; Nadia A. de Vries; Marcel Wittenberg, Jana Riecke.

## Abstract

**Background:** Since the beginning of the coronavirus disease 2019 (COVID-19) pandemic, there has been increasing demand to identify predictors of severe clinical course in patients infected with severe acute respiratory syndrome coronavirus 2 (SARS-CoV-2). Human leukocyte antigen alleles (HLA) have been suggested as potential genetic host factors. We sought to evaluate this hypothesis by conducting an international multicenter study using HLA sequencing with subsequent independent validation.

**Methods:** We analyzed a total of 332 samples. First, we enrolled 233 patients in Germany, Spain, and Switzerland for HLA and whole exome sequencing. Furthermore, we validated our results in a public data set (United States, n=99). Patients older than 18 years presenting with COVID-19 were included, representing the full spectrum of the disease. HLA candidate alleles were identified in the derivation cohort (n=92) and tested in two independent validation cohorts (n=240).

**Results:** We identified HLA-C* 04:01 as a novel genetic predictor for severe clinical course in COVID-19. Carriers of HLA-C* 04:01 had twice the risk of intubation when infected with SARS-CoV-2 (hazard ratio 2.1, adjusted p-value=0.0036). Importantly, these findings were successfully replicated in an independent data set. Furthermore, our findings are biologically plausible, as HLA-C* 04:01 has fewer predicted bindings sites with relevant SARS-CoV-2 peptides as compared to other HLA alleles. Exome sequencing confirmed findings from HLA analysis.

**Conclusions:** HLA-C* 04:01 carriage is associated with a twofold increased risk of intubation in patients infected with SARS-CoV-2. Testing for HLA-C* 04:01 could have clinical implications to identify high-risk patients and individualize management.

## INTRODUCTION

Severe acute respiratory syndrome coronavirus 2 (SARS-CoV-2) was first reported in Wuhan, China, at the end of 2019^1,2^. It rapidly spread to Europe, the United States, and the rest of the world, manifesting as coronavirus disease 2019 (COVID-19)^3,4^.

Since the beginning of the pandemic, a striking clinical spectrum has been described among patients with COVID-19. Some patients remain asymptomatic, others may become “super spreaders” or “super emitters^5^, while yet others have a severe clinical course leading to respiratory or multiorgan failure with potentially lethal outcome^6^. Various clinical risk factors for severe clinical course have been described such as age, diabetes, hypertension, and obesity^4,6^. On the genomic level, it has been suggested that blood groups^7,8^, as well as genes involved in antiviral defense mechanisms and inflammatory organ damage, may affect clinical outcomes^9^. In addition to blood groups, the HLA system plays a crucial role in immune response^10,11^. Genetic polymorphisms of HLA have been reported to affect the clinical course of patients infected with various ribonucleic acid viruses (e.g. H1N1^12^, Hantaan virus^13^, and SARS-CoV- 1^14^).

Therefore, we sought to test the hypothesis that specific HLA alleles predispose patients to a severe clinical course in COVID-19. Severe clinical course was defined as acute respiratory failure, requirement for intensive care, or death. In addition, we sought to evaluate if specific HLA alleles predispose patients to acute cardiac injury.

Data from the SARS-CoV pandemic suggest that this was a plausible hypothesis. HLA-B* 46:01 has been shown to be associated with a severe clinical course in patients infected with SARS-CoV. Furthermore, a recent in silico analysis of HLA data from an international database suggested that HLA-B* 46:01 appears to predispose patients to a more severe clinical course of SARS-CoV-2^15^, similar to that found in actual clinical samples from patients with SARS-CoV-1^14^. While HLA-B* 46:01 frequency is 17% in Asia, where these data were obtained, the allele frequency is 0.05% in Germany and <0.01% in Spain, where the largest part of our study was conducted (*http://www.allelefrequencies.net*)^16^. Therefore, we suspected that if an HLA risk allele was identified, it would differ from the one discovered during the SARS-CoV pandemic in Asia.

To test our hypothesis, we collected blood samples from patients infected with SARS-CoV-2 for HLA sequencing during the first peak of the European COVID-19 pandemic. Enrollment centers included hospitals in Germany, Switzerland, and Spain. Furthermore, we leveraged the extant RNA-Seq data sets available from the Gene Expression Omnibus, which contained samples from the United States to validate our findings.

## METHODS

### Study participants and recruitment

We recruited a total of 233 patients with mild to severe symptoms of COVID-19 for HLA sequencing analysis. In addition, we validated our findings in data from a public data set containing RNA sequencing data for a total of 99 patients with COVID-19 containing phenotypic information on requirement for intensive care, intubation or death *(https://www.ncbi.nlm.nih.gov/geo; GSE157103*^*17*^). Participants in our cohort were representative of the full spectrum of clinical presentation with COVID-19 from asymptomatic to multiorgan failure and death. Diagnosis was made based on detection of SARS-CoV-2 viral RNA using a reverse transcription-polymerase chain reaction (RT-PCR) test from nasopharyngeal swabs and/or detection of SARS-CoV-2 specific antibodies in the blood through enzyme-linked immunosorbent assay (ELISA). Sample collection took place during the first peak of the pandemic at five European Hospitals: Germany: Charite Universitätsmedizin Berlin (3 sites, total 92 patients, data set 1, DS1)^18,19^; Spain: Hospital Universitario de Valme, Sevilla (total 120 patients) and Switzerland: Kantonsspital Baden AG, Baden (total 21 patients, data set 2, DS2). All patients with COVID-19, who provided informed consent and were 18 years or older were included. Whole-blood samples, buffy coats, or peripheral blood mononuclear cells (PBMCs) were collected from all patients for DNA extraction during routine diagnostic venipuncture. In addition to clinical and laboratory data, we had access to first measured viral load data for the cohort from Germany (obtained by RT-PCR)^20^.

We used requirement for intensive care unit (ICU) admission, intubation, or death as well as a combined endpoint of all three outcomes as categories for HLA analysis.

We used the standard reference range for troponin T hs (<14ng/l) to define troponin elevation. Acute cardiac injury was defined as elevation of troponin T hs (>14ng/l) without obvious signs for acute coronary syndrome. Furthermore, we directly used the troponin T hs levels to test for association. Troponin T hs levels were available for 66 patients.

### Ethics committee approval

Our protocol included blood samples obtained during routine clinically indicated venipunctures, which were then frozen at -80°C for subsequent deoxyribonucleic acid (DNA) extraction and sequencing. A minimal set of deidentified clinical data was collected to protect data privacy. At Charite Universitätsmedizin Berlin procedures were performed within the framework of the Pa-COVID-19 protocol^18^.

Given the exceptional situation of the pandemic and the international character of this study, there were differences amongst local ethics committees in consent procedures^7^. At each center, written informed consent was obtained from patients when possible, sometimes retrospectively if the patient was not able to give consent at the moment of sample collection.

This study was approved by the ethics committee of Charité Universitätsmedizin Berlin, Germany (EA2/066/20), the Ethikkommission Nordwest- und Zentralschweiz, Basel, Switzerland (2020-00952), and the Ethics Committee of the Hospital Universitario de Valme, Sevilla Spain (1346-N-16).

Data sets differ in the depth of clinical annotation and sites used different criteria, e.g. for admitting patients to the ICU.

## DNA extraction, HLA sequencing, and quality control

### DNA extraction

DNA was extracted either manually using the Quick-DNA™ Miniprep Plus Kit (Zymogen, number D4068) or automatically on a Roche LC2 robot according to the manufacturer’s instructions. On the robot The MagNA Pure LC DNA Isolation Kit - Large Volume (Roche, number 03310515001) was used. DNA concentration was measured using either Nanodrop (Thermo Fisher) for the Zymogen extracted samples and Qubit™ dsDNA HS Assay Kit (Thermo Fisher, number Q32851) for robot-extracted samples.

### HLA typing by PacBio SMRT sequencing, PCR Amplification of Full-Length HLA Genes

Aliquots of patient genomic DNA samples were diluted to the same concentrations in 96-well plates. To meet the challenges of a high-throughput project, further pipetting steps were performed with multichannel pipettes or on the epMotion 5075 pipetting robot (Eppendorf).

For each patient sample, eleven HLA genes were amplified using NGSgo®-AmpX v2 HLAGeneSuite kit (#7371662, GenDx). According to the design of this protocol, amplicons cover all class I HLA genes (HLA-A, -B, -C) and the most relevant parts of the class II HLA genes (HLA-DRB* 1, -DRB* 3/4/5, -DQB* 1, -DPA* 1, -DPB* 1, -DQA* 1), which have very long introns. Amplification was performed according to the manufacturer’s instructions *(https://www.gendx.com/product_line/ngsgo-ampx-v2).* To improve PCR efficiency, the starting amount of template DNA was increased from 45ng to 80ng per reaction for loci HLA-DRB* 1, - DQB* 1, -DPA* 1, -DPB* 1 and –DQA* 1. In addition, elongation time for HLA-DQA* 1 was extended to 5min. Nevertheless, assay performance differed between the loci. Amplification was typically unproblematic for HLA-A, -B, -C, and DQA1 genes and less consistent in terms of output for other loci.

Amplicons were purified with Agencourt AMPure XP beads (#A63881, Beckman Coulter). Quality and quantity of purified PCR amplicons were checked using D5000 Screen Tape assay (#5067- 5593, Agilent) on TapeStation (Agilent) and Qubit dsDNA assay kit (#33120, Thermofisher) on FLUOStar Omega microplate reader (BMG Labtech), respectively. Confirmed amplicons were equimolar combined into two– for HLA class I and HLA class II – pools per patient.

### HLA SMRTbell Library Preparation and Sequencing

Single molecule real time (SMRT) sequencing libraries were prepared according to the manufacturer’s protocol *(Procedure & Checklist -Preparing SMRTbell™ Libraries using PacBio® Barcoded Adapters for Multiplex SMRT® Sequencing PacBio®)*^*21*^. Barcoded Adapters were used to label patient amplicon pools. Up to 72 amplicon pools were combined per HLA class I and HLA class II libraries. Libraries were validated using 12000 DNA Assay (#50697-1508, Agilent) on Agilent Bioanalyzer and quantified using Qubit dsDNA assay on Qubit Fluorometer.

Libraries were loaded on SMRT cells and sequenced on Sequel instrument (Pacific Biosciences) using 10h / SMRT Cell 1M sequencing runtime. Loading setup and run design was planned with SMRTLink v9.0 *(https://www.pacb.com/wp-content/uploads/Quick-Reference-Card-Loading-and-Pre-Extension-Recommendations-for-the-Sequel-System.pdf)*. In total, HLA genes amplicons for 229 patients were sequenced on 12 SMRT cells.

Circular consensus sequence (CCS) data analysis and demultiplexing was performed within SMRT Link 9.0.0.92188 GUI.

### Validation with RNA-Seq data (data set 3, DS 3)

Publicly available data were downloaded from the Gene Expression Omnibus (GEO identifier GSE157103) and analyzed using the arcasHLA package, version 0.2.0^21^.

Ulitmately, correlation analysis was performed to compare HLA allele frequencies between all three data sets.

### Exome sequencing

Genomic DNAs were quantified by Quant-iT™ dsDNA Assay Kit, broad range (ThermoFisher Scientific) with a FLUOStar Omega microplate reader (BMG Labtech). Illumina-compatible libraries were prepared on a Bravo NGS Workstation Option B according to the manual KAPA HyperPrep/HyperPlus Automated Workflow for Agilent Bravo B NGS Instructions for Use, v3.0 (Roche Sequencing) using the KAPA Hyper Plus workflow with the reagents included in the KAPA Hyper Plus kit (Roche Sequencing). In short, 100 ng of genomic DNA was fragmented at 37°C in an Eppendorf Mastercycler Plus S for 25 minutes with subsequent end repair. Following end repair, fragments were ligated to the KAPA Universal Adapter (Roche Sequencing) and purified by KAPA HyperPure Beads (Roche Sequencing). Purified libraries were amplified by 6 cycles of PCR utilizing the KAPA UDI Primer Mix* (Roche Sequencing) and again purified using KAPA HyperPure Beads*. Amplified sample library concentration and size distribution was determined by Qubit dsDNA HS assay kit (ThermoFisher Scientific) with a Qubit™ 3 Fluorometer (ThermoFisher Scientific) and D5000 ScreenTape assay (Agilent Technologies) with a 4200 TapeStation system (Agilent Technologies). Eight amplified sample libraries were pooled prior to enrichment. For this purpose, 187,5 ng of each library was pooled into one well and the final volume was adjusted to 45 µl with nuclease-free water as designated by the manual KAPA HyperCap Workflow v3.0 (*https://pim-eservices.roche.com/eLD/web/pi/en/home*) and all further steps were performed following the recommendations of that manual.

These included hybridization of the amplified sample library pool to KAPA HyperExome Probes (Roche Sequencing) washing and recovery of the captured multiplex sample library, as well as amplification and purification of the enriched multiplex sample library. Final library pools were quantified by Qubit dsDNA HS assay kit with a Qubit™ 3 Fluorometer and correct size distribution was validated by High Sensitivity D5000 ScreenTape assay (Agilent Technologies) with a 4200 TapeStation system. A total of 12 multiplex enriched sample libraries were pooled equimolar utilizing the NextGen HyperCap Pooling Guide and Calculator template (Roche Sequencing) and diluted to 1.3 nM. Each final pool was sequenced on two lanes of an S4 FlowCell (Illumina, Inc.) for 2x 151 cycles on a NovaSeq 6000 system (Illumina, Inc.) *For Research Use Only. Not for use in diagnostic procedures.

### Exome analysis

Exome data was aligned using BWA-MEM to the reference GRCh37 (hs357d.fa). Samples were identified as being KIR2DS4f positive by screening the GATK HaplotypeCaller v3.7^22^ variant calls for presence of an insertion of CCCGGAGCTCCTATGACATGTA in exon 4 of KIR2DS4. For a more detailed description of the analysis please see methods in the data supplement.

## Statistical analysis

### Composite disease score

DS1 consisted of patients of the Charite cohort. For patients in DS1, we defined a composite score, including troponin T hs levels, death, ICU and intubation status (binned in five categories). Then, we used multiple correspondence analysis (MCA; R package FactoMineR^23^, v. 2.3) to obtain a score derived from the selected variables, defined as the first dimension of the individual coordinates.

### HLA association testing

Dominant model of association with the selected response variables was tested using Fisher’s exact test (for categorical variables) or t-test (for continuous variables). The logarithm of Troponin T hs levels was used for the association test. In the analysis, we only considered alleles with a frequency in the given data set above 5% and with at least 5 individual carriers of the allele present. Other alleles were retained for the purpose of association testing, but not directly evaluated for association. The response variables differed between the three data sets analyzed. DS 1 (Berlin) included the following continuous variables: Troponin T hs, maximum WHO ordinal scale for clinical improvement score, maximum troponin T hs levels, and viral load, as well as categorical: Troponin T hs elevation above 14ng/l, ICU and intubation status, and death. For Data Sets 2 and 3 (Switzerland/Spain and US, respectively; DS 2 and 3) we tested for association with ICU and intubation status (categorical). To calculate the effect size, we have used Cramer’s v for categorical variables and Cohen’s d for continuous variables.

### Randomization test

To obtain an empirical p-value of our selection / validation process, we have used a randomization test. We replicated the process 10,000 times, each time randomly assigning the real genotypes within a given data set to the patient identifiers. Then, for each allele, we calculated how many times the best p-values obtained in each of the three data set for associations with the different response variables within the randomization test are all lower than the p-values obtained with the non-randomized data set. The p-values obtained with randomization test were then recalculated using the Benjamini-Hochberg procedure to control for false discovery rate (FDR)^24^.

### Genetic structure of DS1 and DS2

To test presence of subpopulations in DS1 and DS2, we leveraged exome derived variants. We used the R package SNPrelate, version 1.2.1, to process the VCF file and perform a principal components analysis. Next, we tested whether any of the principal components were associated with either intubation or ICU status or with the presence / absence of the identified risk allele.

## RESULTS

### Patients

Descriptive data on age, sex, intensive care management, maximum respiratory support during hospitalization and relevant comorbidities (hypertension, coronary artery disease, diabetes) of patients included in the primary and second data set are illustrated in table 1.

**Table 1.**
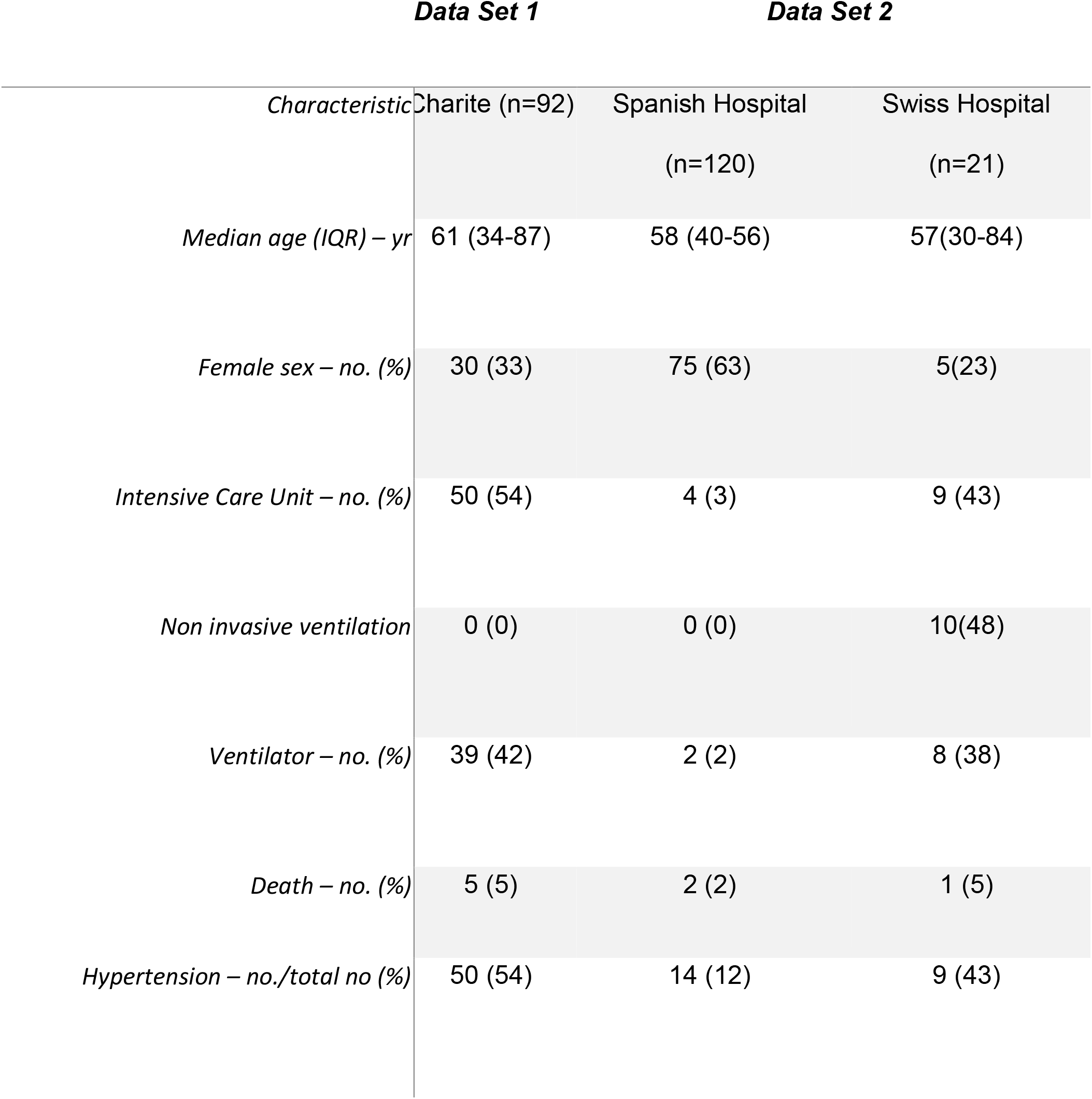
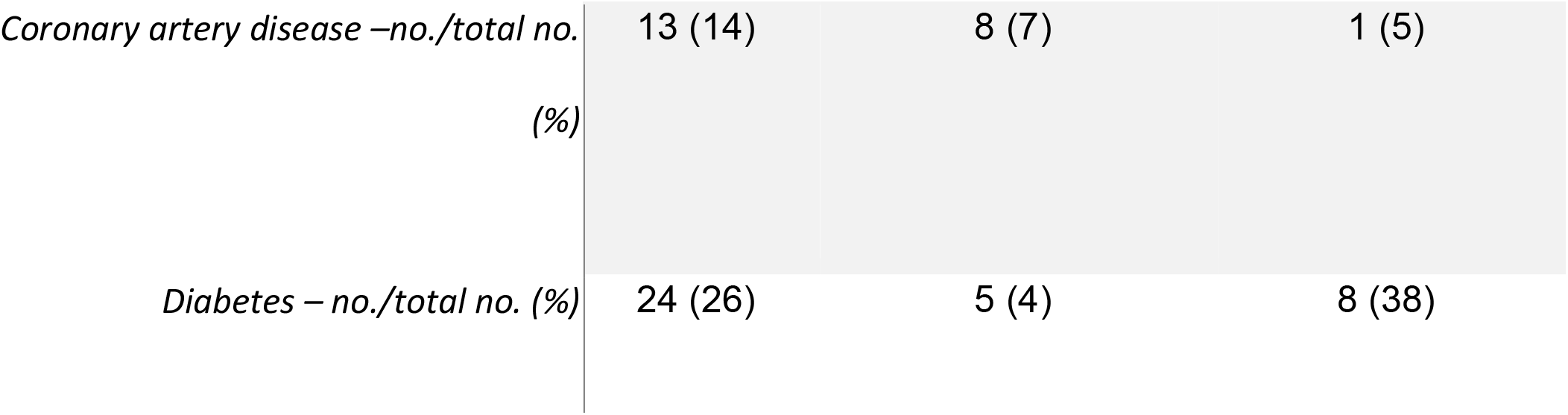
Clinical parameters of patients in data set 1 and 2

The median age was similar in the derivation cohort (DS1) and the two validation cohorts (DS 2 and 3), ranging from 57 to 61 years. The Spanish cohort consisted of more women (63%) than the German (33%) and the Swiss (24%) cohorts. In the Spanish cohort the prevalence of hypertension and diabetes was lower and patients were less likely to get intubated.

The genetic structures of DS1 and DS2 showed no indication of existing subpopulations based on principal component analysis of variants derived from whole exome sequencing data. None of the components was significantly associated with ICU or intubation status, even though few individuals were of different genetic background.

### Stepwise analysis of HLA associations

Given the relatively small, but well-annotated DS1, we decided on a step-wise validation approach (Figure 1). In essence, we used DS1 as a discovery cohort to propose candidate alleles which then were validated using DS 2 (Switzerland/Spain) and DS 3 (United States). Calculation of overall associations on a combined data set of all three cohorts (see Supplementary table 1) is prone to discovery of false positives, due to varying allele frequencies and percentages of apparently severe disease cases in the three data sets. To avoid this, as well as other potentially incorrect assumptions regarding variable distribution, we decided to test the obtained results using a randomization approach. In short, we repeated the whole selection / validation procedure 10,000 times, each time randomly assigning the obtained HLA genotypes to patients (separately in each cohort) and used the smallest p-values for associations in each data set as our test statistics. Such a randomization test allowed us to estimate how often the obtained combination of p-values from discovery and validation cohort occurs when there is no real association between alleles and response variables.

**Figure 1.**
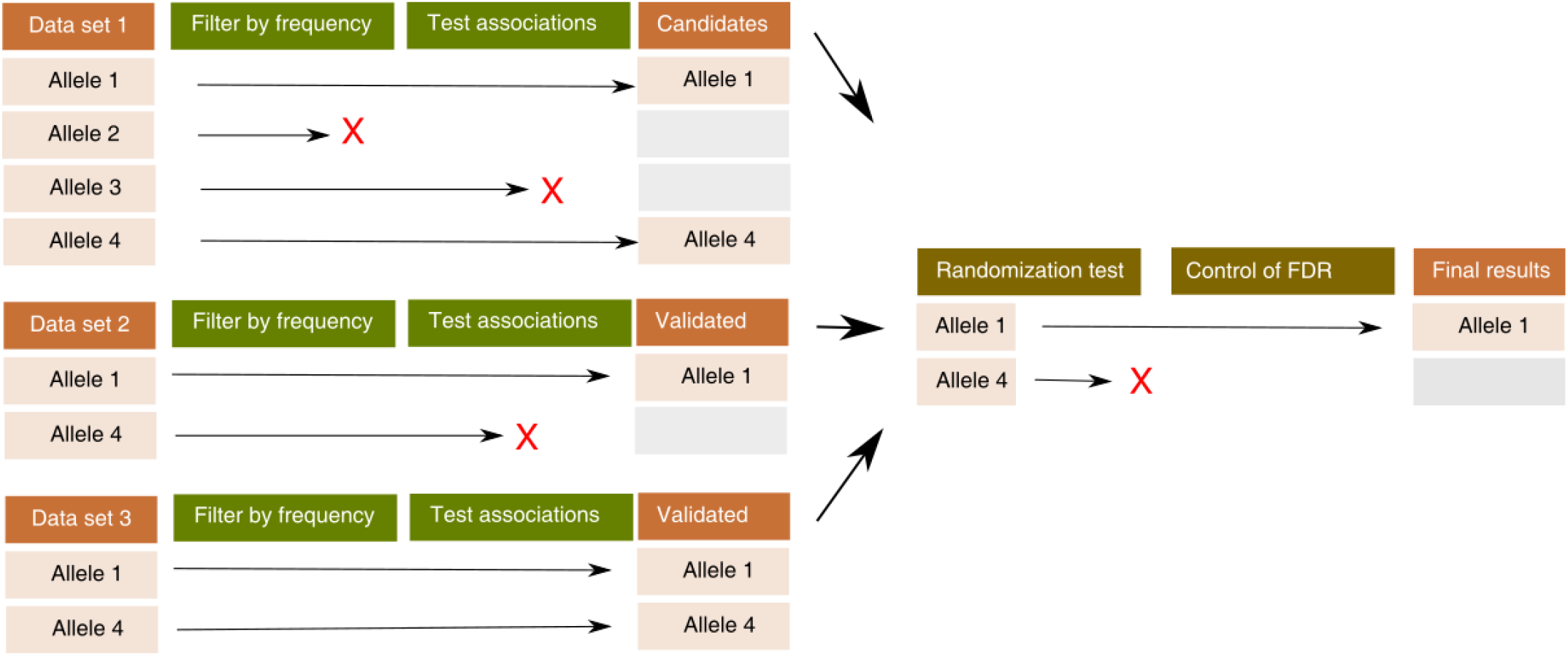
Scheme of the discovery and validation of HLA alleles associated with COVID-19 severity. First, all alleles found in DS1 are filtered by frequency and tested for association with a number of variables considered to reflect the disease severity, such as ICU or intubation status. Then, the allele candidates are tested for association with ICU and intubation status in DS2 and DS3. Finally, the whole procedure is repeated 10,000 times with genotypes randomly associated with the patient data (randomization test) and the obtained p-values are corrected for false discovery rate (FDR).

### Candidate alleles from DS1

For selecting candidates, we defined a cutoff allele frequency of >5% and a minimum number of carriers (5) in the study population to ensure the relevance of the potential findings. Next, we tested the associations of the remaining alleles in DS1 after filtering with each of the proposed response variables considered to be proxies of disease severity: ICU and intubation status, troponin T hs levels, maximum WHO ordinal scale for clinical improvement score and a composite score. We selected alleles as candidate alleles, if they showed at least one association at p < 0.05 (not adjusted for multiple testing). We found 16 significant associations of 11 alleles (Figure 2; Supplemental Table 2); the association between allele HLA C*03:03 and the composite score was significant after correcting for FDR.

**Figure 2.**
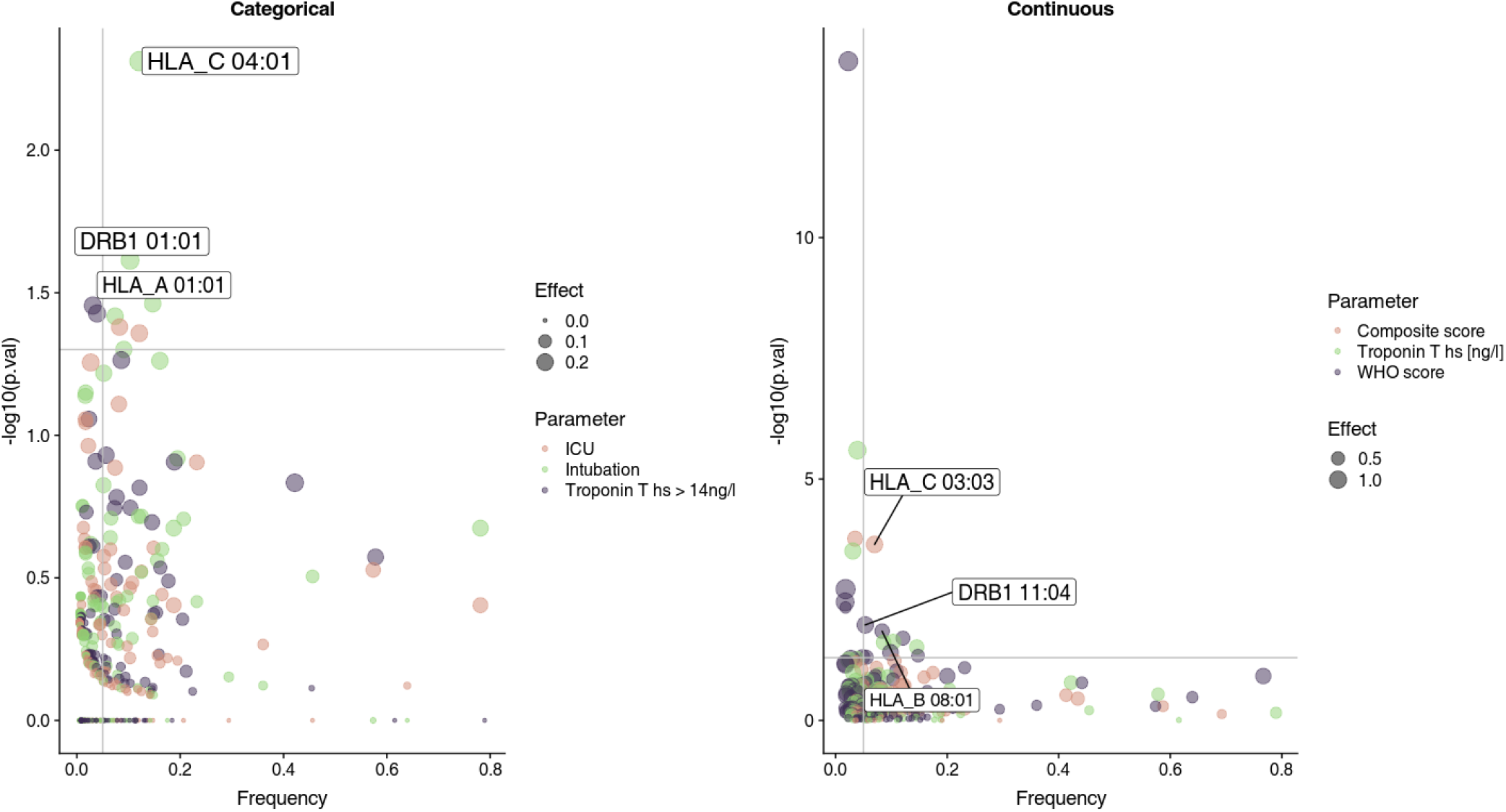
Allele candidate selection by testing associations in DS1. Left, associations with categorical variables; right, associations with continuous variables. Each dot represents one association between a variable (e.g. intubation status) and an allele (e.g. HLA C*04:01). Dot color indicates the response variable tested; vertical dot position indicates the p-value, dot size indicates the effect size (Cramer’s v for categorical variables, Cohen’s d for continuous variables), while horizontal position indicates the frequency of the allele. Grey vertical and horizontal lines show candidate selection criteria: p < 0.05 (horizontal line) and frequency > 0.05 (vertical line). For the three top associations (by p-value), names of the alleles are shown as labels.

Overall, the most frequent significant (p < 0.05) associations were with maximum WHO ordinal scale for clinical improvement score, intubation status, and troponin, while there were only two associations with ICU status and one with composite score.

Several of the test results indicated that the allele candidates were associated with greater disease severity. For example, while on average one third of the patients were intubated, 15 out of the 21 patients who carried the HLA-C* 04:01 allele were intubated. However, there were also alleles associated with milder disease course. The type II allele DQA1 *05:01 (present in 10 patients) was associated with lower troponin T hs levels; the common type I allele HLA-A* 01:01 (present in 25 patients) was weakly associated with lower WHO ordinal scale for clinical improvement score and lower fraction of intubated patients (24 vs 49%).

### Validation in DS 2 and DS 3

For DS 2 and DS 3, two response variables were generally available for all samples: ICU and intubation status. We therefore tested the association with ICU and intubation status for all candidate alleles. Furthermore, we tested association with troponin T hs levels in the Swiss cohort in DS2, for which this information was available. To determine whether a confirmation in one or both of DS2 and DS3 truly validates the allele candidates from DS1, we have used a randomization test, in which the selection/validation procedure was mimicked using resampled genotypes.

In DS2, there was a significant (at FDR < 0.05) association between the allele HLA-DRB1* 11:04 and intubation status. Notably, amongst the 137 patients of DS 2, only 13 were treated in an ICU and 10 were intubated. Out of the 10 patients who were intubated, three were carrying the HLA- DRB1* 11:04 allele. This association was not confirmed by the randomization test. In DS3, the only significant associations were between the allele HLA- C* 04:01 and both ICU and intubation status (Figure 3; Supplemental Figure 1). The p-value obtained from the randomization test for this allele was < 0.004 after correcting for FDR (Supplemental Table 3), even though no association between HLA-C*04:01 could be detected in DS 2.

**Figure 3.**
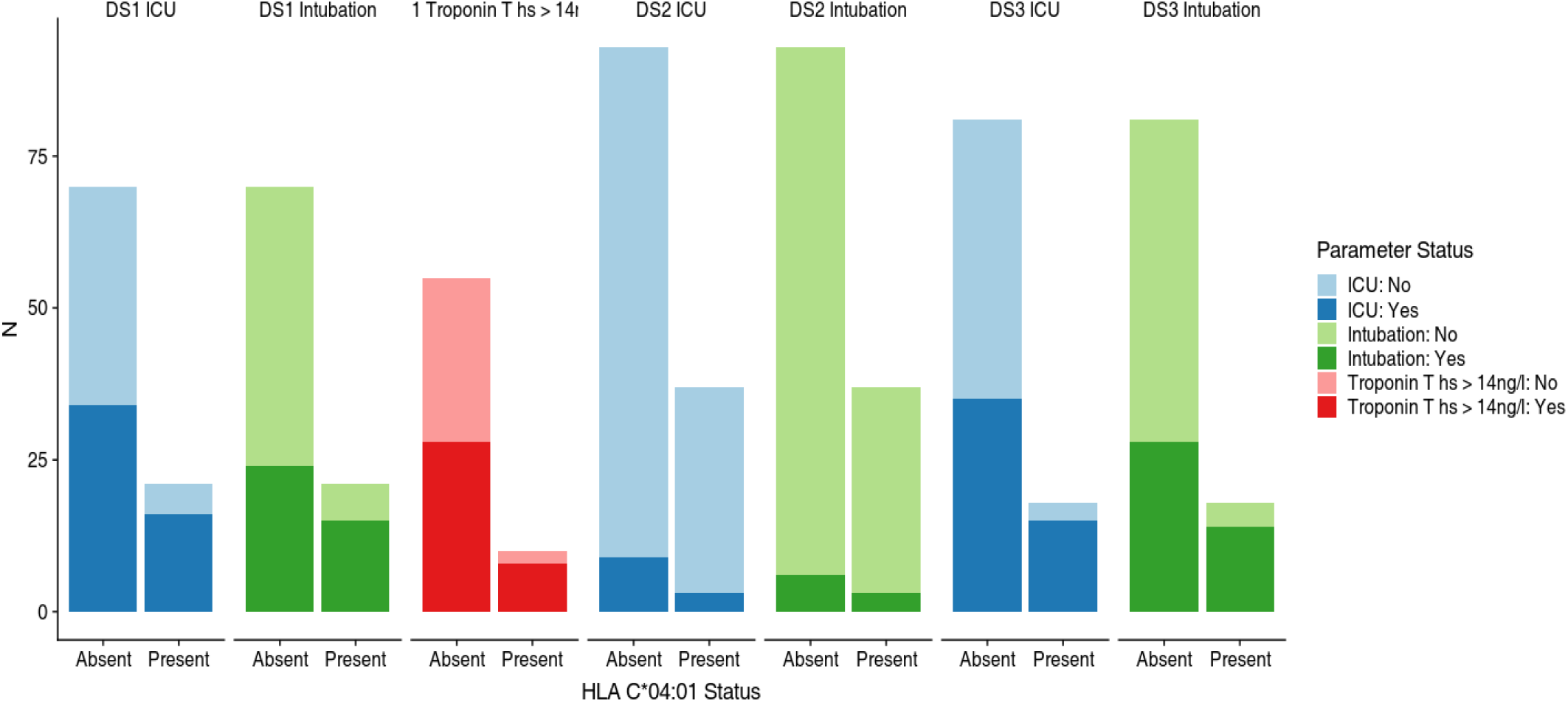
Associations of HLA C*04:01 with categorical response variables. Each panel shows the difference between patients who were carrying the allele (HLA C*04:01 status: Present) and patients who did not carry the allele (HLA C*04:01 status: Absent). Colors correspond to variable status. Vertical axis shows the absolute number of patients. D1 - DS3: data sets 1 - 3.

Finally, correlation analysis of allele frequencies overall revealed that HLA alleles had comparable frequencies between Germany, Switzerland, Spain, and the United States (Supplemental Figure 2).

### Association of alleles with viral load

For DS1, first measured viral load data were available. Notably, measured viral load was not associated with severity of symptoms such as ICU/intubation status or troponine levels (Supplemental Figure 3, Supplemental Table 1). There were five significant associations of initial viral load with HLA alleles at p<0.05, none of which were significant after correction for false discovery rate. The HLA-C* 04:01 allele was not significantly associated viral load. Given that viral load was not associated with disease severity in DS1 and that viral load data were not available for DS2 or DS3, we did not attempt to confirm whether the lack of association between HLA and viral load in DS1 was mirrored in the two other datasets.

### Association between HLAs and other covariates in DS 1

One possible explanation of the association between HLA alleles and COVID-19 severity is that these alleles occur at different frequencies in different risk groups. Therefore, we sought to evaluate if HLA-C* 04:01 occurs with higher frequency amongst older patients, who are at greater risk of developing severe symptoms of COVID-19. Indeed, patients who were intubated or admitted to the ICU were older overall (p-values from a two-sided t-test, were 0.003 and 0.014, respectively). However, HLA-C* 04:01 had an overall frequency of 0.12 and 0.14 in patients younger than 65 and a lower frequency, 0.067, in the high-risk group of patients older than 75 years.

To exclude other potential confounding effects, we tested the associations of all alleles in DS 1 with the covariates gender, age, weight, and body mass index (BMI). While there were seven alleles which showed a significant association at p < 0.05 (Supplemental Table 5), no association was significant after correction for multiple testing.

Furthermore, we tested if HLA-C* 04:01 is associated with higher CRP levels as a surrogate for systemic inflammation. However, there was only a trend that was not significant (Supplemental Figure 4). CRP correlated with the risk of intubation.

### Affinity analysis

A possible explanation for the effect of an allele on disease severity might be abnormal binding affinity to SARS-CoV-2 peptides. To test that hypothesis, we used a previously described data set of putative (computed in silico) binding affinities.^15^ Following an approach described by Iturrieta-Zuazo and colleagues^25^, we calculated the number of SARS-CoV-2 peptides, which bind either “tightly” (at < 50 mM) or “loosely” (at < 500 mM) for each allele. Using this approach, we discovered that HLA C*04:01 is among the ten alleles with the fewest predicted binding SARS-CoV-2 peptides (Figure 4). Therefore, we sought to systematically test the hypothesis that the putative affinity to SARS-CoV-2 peptides is a correlate of disease severity. First, for each of the response variables (ICU and intubation status, troponin T hs levels, composite score) we tested whether the effect size is correlated with the number of potentially-binding peptides. We included all alleles (not filtered for frequency) in this comparison. While in all data sets combined there were no large effects in alleles with many potential binding peptides, negative correlation was only significant in DS3 (Spearman test, p < 0.007, *ρ*=0.27).

**Figure 4.**
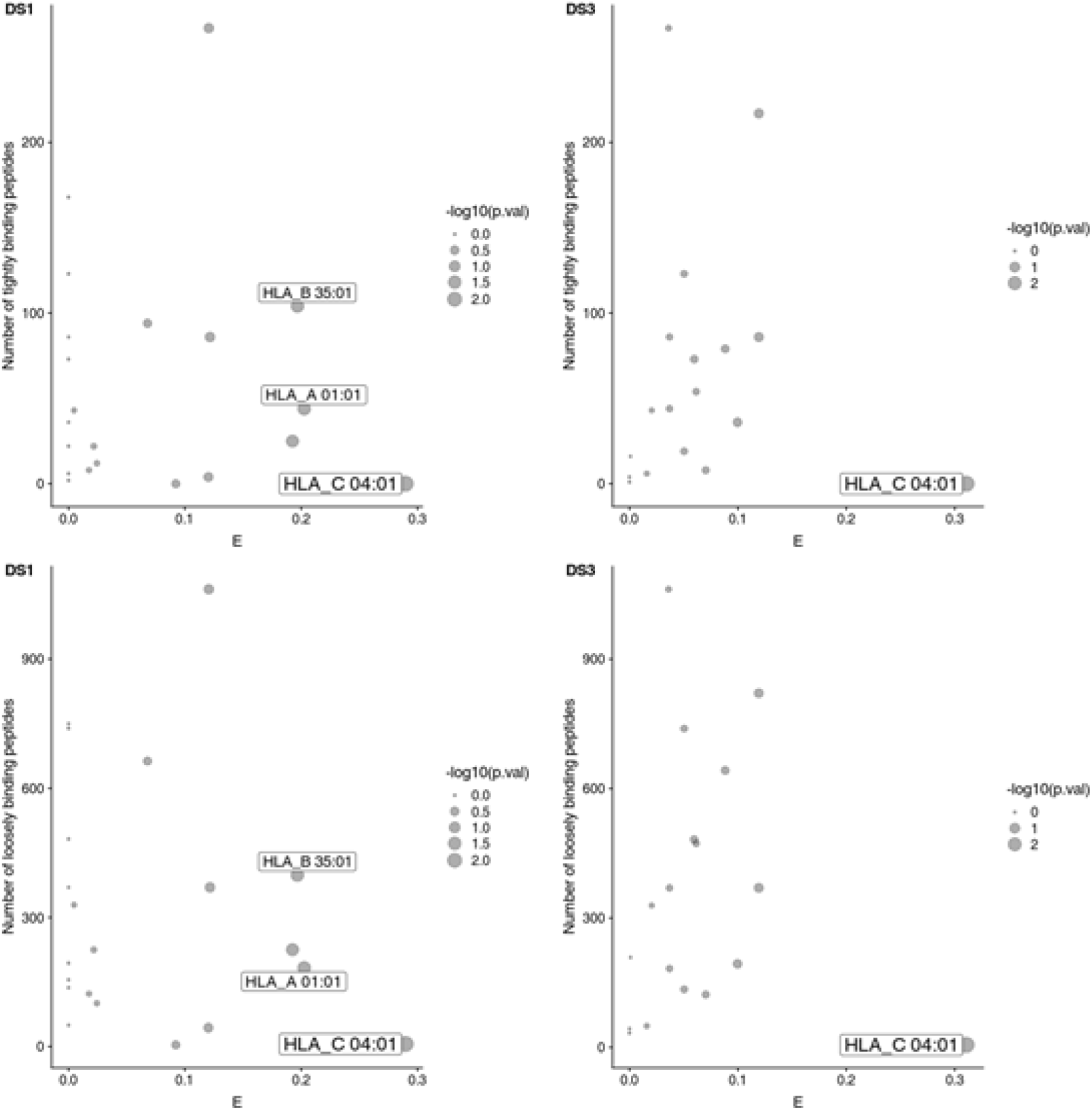
Correlation between the estimated effect size for the association between alleles and intubation status and the number of peptides putatively recognized by that allele. Each dot corresponds to one allele. Left column shows plots for DS1, right column shows plots for DS3; upper row shows plots for number of “tightly” binding peptides, lower row shows plots for number of “loosely” binding peptides. Dot size corresponds to the p-value in Fisher’s exact test. Alleles showing significant associations with intubation status are labeled.

Next, for each patient, we calculated how many SARS-CoV-2 peptides can the type I alleles in that patient potentially bind, and tested whether there is an association between disease severity (using troponin T hs levels, intubation and ICU status and composite score as proxies) and that number. However, none of these tests showed a significant association.

### Exome sequencing data

Since polymorphisms in the KIR2DS4 gene in combination with the HLA*C 04:01 allele have been reported to increase viral load and lead to severe clinical course in patients with human immunodeficiency virus (HIV)^26^, we evaluated if presence of the KIR2DS4f variant was associated with our outcome variables (e.g. viral load, intubation). However, no association was identified. A combination of a KIR2DS4f variant with HLA*C 04:01 was identified in only four patients, of which only one was homozygous at the KIR2DS4 locus. This patient had a severe clinical course, was intubated, and had highly elevated troponin T hs and maximum WHO ordinal scale for clinical improvement score.

## DISCUSSION

In the beginning of the outbreak of the COVID-19 pandemic in Europe in spring 2020, an international collaboration of European centers was established to address the question, whether there were potential genetic host factors associated with severe clinical course in SARS-CoV-2. Samples were collected in Spain, Switzerland, and Germany^18^ and subsequently processed in Germany for full-length sequencing of HLA genes in DNA samples.

We identified HLA-C* 04:01 as the most relevant risk allele, which was associated with a twofold risk of intubation when present in the form of at least one allele.

Importantly these findings were reproduced in an independent public data set obtained from patients with COVID-19 at Albany Medical Center, United States using RNA sequencing^17^. Using this approach, we demonstrated broad applicability of our findings and confirmed reproducibility and robustness of our results in data gathered through a distinct method – the analysis of the transcriptome^27^. This is particularly relevant in the context of regional differences in allele frequencies^28,29^. In that regard, HLA-B* 46:01 was shown to be associated with severe clinical course in SARS-CoV in Asia^14^. However, this allele is very rare in Germany, Spain, Switzerland and the United States^16^. In line with that, HLA-B* 46:01 was not represented in our European data set. In a correlation analysis we demonstrated that the frequencies of most alleles of the German cohort were similar to those in the cohorts from Switzerland, Spain, and the United States.

After completing our stepwise analysis of data sets, final randomization and multiple comparisons of all three data sets confirmed that HLA-C* 04:01 was strongly associated with requirement of intubation. While this allele was not found to be significantly associated with disease severity in DS2, this data set included only a small number of patients with severe clinical course (13/133 patients treated at the ICU; 10/133 intubated, figure 3). Therefore, detecting an existing association of HLA-C*04:01 was unlikely due to low statistical power, considering the observed effect size and allele frequency. Nonetheless, the combined effects observed in DS1 and DS3 were sufficient to reach a robust conclusion during randomization. Another factor in which DS 2 differed from DS1, in particular the Spanish population, was that it consisted of a larger percentage of women (63% in Spain vs 33% in Germany). Importantly, DS2 was a representative sample of patients in Sevilla, where our recruiting center collected samples consecutively at the peak of the pandemic and by now has seen a similar proportion of women (60%) in all patients, who have been recruited since then (n=842). While our analysis did not reveal any association of specific HLA alleles with sex (supplemental table 5), it has been shown that female sex is associated with better outcomes^30^. This effect of sex may have masked the effect of HLA-C*04-01 validation in DS 2. Future research is necessary to investigate this hypothesis.

In addition to the strong and robust association of HLA-C* 04:01 and intubation with a hazard ratio of 2.1, there is also evidence in the literature suggesting a potential biological role of HLA- C* 04:01 in the defense of viral infections. In that regard, it has been shown that in patients infected with the human immunodeficiency virus (HIV), HLA-C* 04:01 in combination with polymorphisms in the KIR2DS4 gene was associated with high viral titers and severe clinical course^26^. Moreover, HLA-C* 04:01 has been reported to occur at a higher frequency in COVID- 19 patients than in the healthy population^31^.

The allele frequency of HLA-C* 04:01 is approximately 13% in Germany overall and 19% in German residents of Turkish origin^16^, suggesting that the presented findings may impact a relevant fraction of the German population. Similar numbers are found in Switzerland, where the allele frequency is about 16% and Spain, where an allele frequency of 15% has been reported^16^.

On the broader scale, it is notable that the highest allele frequencies of HLA-C* 04:01 worldwide have been observed in Oaxaca, Mexico (38%); Baloch, Iran (29%); Australian Aborigines (28%); and African Americans (21%), while countries such as Japan and South Africa have very low frequency (4% each)^16^. While socioeconomic and classical risk factors, such as population age and comorbidities, are most likely the strongest predictors for severe clinical outcomes in patients with COVID-19, our data raise speculation that disease severity may be exacerbated in populations with high HLA-C* 04:01 frequency.

Since HLA-C* 04:01 is associated with viral load in HIV, we sought to evaluate if this was true for Sars-CoV-2. We did not find any association of HLA-C* 04:01 with first measured viral load of patients during their hospitalization. In addition, the first measured viral load in intubated patients did not differ from patients with mild disease. We support the common hypothesis that by the time the patients present with severe respiratory failure, the virus may have been already partially cleared from the body in some of them and the disease may be maintained through inflammatory or autoimmune mechanisms similar to other viral respiratory illnesses.^32^ The dynamics of viral load in SARS-CoV 2 have been reported comprehensively in the literature^20,33,34^. It may be beneficial to test for HLA-C* 04:01 in clinical trials to identify from which drug class this high-risk population benefits most.

It is also possible that viral load of patients with HLA-C* 04:01 may have been higher during the very first days of infection^35^, but since most patients only become symptomatic after approximately a week^36,37^, this time window may have been missed in some patients of our cohort to see a potential relationship of HLA-C* 04:01 and viral loads measured during early infection. In general, there was no association of any specific HLA allele with first measured viral load in our cohort.

Exome sequencing revealed a combination of a KIR2DS4f variant with HLA*C 04:01 in only four patients. One of those patients was homozygous at the KIR2DS4 locus and had a severe clinical course. While these findings are interesting, the sample sizes of these subgroups are too small to draw any conclusions.

Notably, HLA affinity analysis revealed that HLA-C* 04:01 binding affinity to SARS-CoV-2 peptides was amongst the ten lowest HLA alleles. The effect size for the association between the allele and intubation status was strong. Delayed immune response due to low HLA binding affinity may be one potential biological explanation for the severe clinical course observed in patients with HLA-C* 04:01.

Alternatively, HLA-C* 04:01 may have led to worse outcomes by predisposing patients to a cytokine storm. To test that, we evaluated for a potential association of HLA-C* 04:01 with CRP serum levels as a surrogate for cytokine storm. Indeed there was a trend of higher CRP levels in HLA-C* 04:01, albeit this finding was not significant. CRP correlated with the risk of intubation, as shown in prior studies^38,39^.

HLA-DQA1* 05:01, HLA-B* 08:01, and HLA DPB* 02:01 were associated with higher troponin T hs levels in DS1. However, this effect was not reproduced in the final validation in DS2 and DS3.

A major strength of this study was the execution of full-length sequencing of HLA alleles, as phasing of single nucleotide polymorphisms (SNPs) is frequently not feasible with the short reads typically used in full genome sequencing^40^. In that regard, any two alleles that differ only at regions that are further apart than the longest read length of the method used cannot be phased, resulting in ambiguity of HLA typing or miscalled alleles^40^. In addition, certain HLA regions, in particular structural variants and insertions, can only be covered with long reads as used in this study. We have confirmed findings from HLA full-length sequencing with HLA calls derived data from full exome sequencing data.

Our study is an important addition to prior HLA research in the field of COVID-19 that provided cumulative evidence for a potential predisposing susceptibility of HLA alleles for infection with COVID-19. It has been previously shown that specific HLA alleles were more common in patients infected with COVID-19 as compared to healthy individuals (e.g. HLA-DRB1* 15:01, HLA-DQB1* 06:02, HLA-DRB1*08, HLA-C*04:01) and it was extrapolated that these alleles may predispose to infection^31,41-43^. It is not surprising that some of these alleles differed from ours, as we identified a genetic predictor specifically for severe clinical course amongst patients with COVID-19, while other groups compared healthy vs diseased. A recent genome wide association study (GWAS) identified blood group A as a major risk factor for COVID-19, but did not detect associations of the classical HLA loci with either COVID-19 infection or disease severity^7^. This study used target short read sequencing of HLA regions for a sub-cohort and investigated association with HLA C:04 only on a 2-digit rather than on the 4-digit level used in this study. Both factors may contribute to a reduced power that made it impossible to detect the association that our approach finds supported in three data sets from different studies and in different populations.

In addition to host factors, it has been shown that genetic factors of the virus may affect affinity of host HLA to viral structures. In that regard, de Sousa and colleagues have shown that a single mutation in the wild type sequence of SARS-CoV-2 could affect peptide binding of the virus to HLA Class II alleles^44^.

In summary, we applied full-length HLA sequencing to patients with COVID-19 to enable identification of HLA alleles with high precision in a large international data set and validated our findings successfully in an entirely independent cohort. We used analysis of clinical and laboratory parameters, viral load data, and whole exome sequencing to evaluate for biological plausibility.

A limitation of this study was that in order to promptly respond to the COVID-19 pandemic with an international multi-center study, even patients with limited metadata were enrolled in this study. However, risk factors and clinical outcomes that we considered most relevant based on prior literature were collected and analyzed for all patients included in our cohorts. Reproducibility of our data in an entirely independent cohort in Albany, United States supports the robustness of our data despite this limitation.

## Supporting information

Cover Letter

## Data Availability

For data privacy reasons, the raw data cannot be made available.

## ACKNOWLEDGEMENTS

We thank all patients who participated in this study for giving us a chance to better understand this novel illness. We would like to express our condolences to the families and friends of patients who died from COVID-19.

We thank the clinical teams at all our enrollment sites for supporting this project while fulfilling their clinical duties during this challenging time of the COVID-19 outbreak.

We thank Gary R. Pasternack, MD, PhD for editorial assistance.

We thank Anja Blüher und Sandra Sieman for excellent technical support.

Biobanking was supported by the Central Biobank Charite.

This work was supported through research funding from Roche Sequencing Solutions, Inc. and a project grant from the Swiss National Science Foundation issued to Bettina Heidecker, MD (Money Follows Researcher Program). This work was supported by the Berlin Institutes of Health (B.I.H. support to the PA-COVID-19 study group, represented by L.E.S and M.W.).

## DATA SUPPLEMENT

### SUPPLEMENTAL METHODS

#### Primary Exome Data Analysis

Exome base call data was converted to FASTQ using bcl2fastq v2.20.0.422. Reads were aligned using BWA-MEM v0.7.15^45^ to the reference GRCh37 (hs37d5.fa), separate read groups were assigned for all reads from one lane, and duplicates were masked using Samblaster v0.1.24.^46^ Standard QC was performed using FastQC.^47^ The variants were then called using GATK HaplotypeCaller v3.7.^22^

#### Secondary Exome Data Analysis: HLA Typing

Exome data was analyzed using OptiType v1.3.5.^48^

#### Secondary Exome Data Analysis: KIR2DS4f Typing

Unlike HLA, no ready-made bioinformatics tool is available for accurately haplotyping the genes in the KIR family, only KPI^49^, which requires whole genome sequencing data. We thus limited the analysis of the functional variant of KIR2DS4^50^ being present (KIR2DS4f+) in the data as follows: The non-functional variant is the major allele of KIR2DS4 that is present in the human reference GRCh37. KIR2DS4f positive samples thus show an insertion of CCCGGAGCTCCTATGACATGTA in exon 4 of KIR2DS4 (dbSNP rs138504928). Therefore, we screened the variant calling results for this variant being present in the data and used the genotype called by GATK HaplotypeCaller for subsequent analysis (0/0 = KIR2DS4f negative, 0/1 = KIR2DS4f het. positive, 1/1 = KIR2DS4f hom. positive).

## SUPPLEMENTAL FIGURES

**Supplemental Figure 1.**
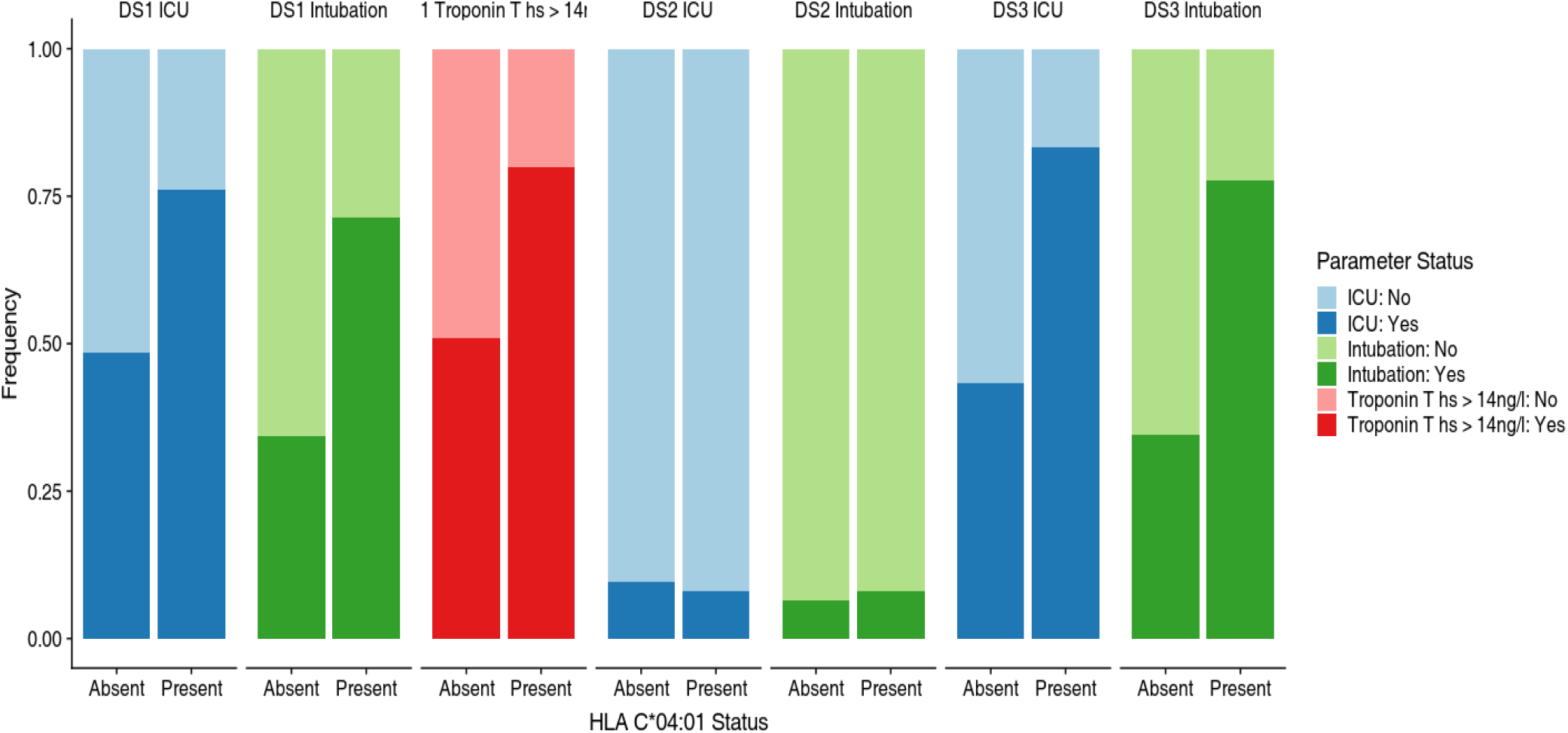
Associations of HLA C*04:01 with categorical response variables. Each panel shows the difference between patients who were carrying the allele (HLA C*04:01 status: Present) and patients who did not carry the allele (HLA C*04:01 status: Absent). Colors correspond to variable status. Vertical axis shows the frequencies of patients. D1 - DS3: data sets 1 - 3.

**Supplemental Figure 2.**
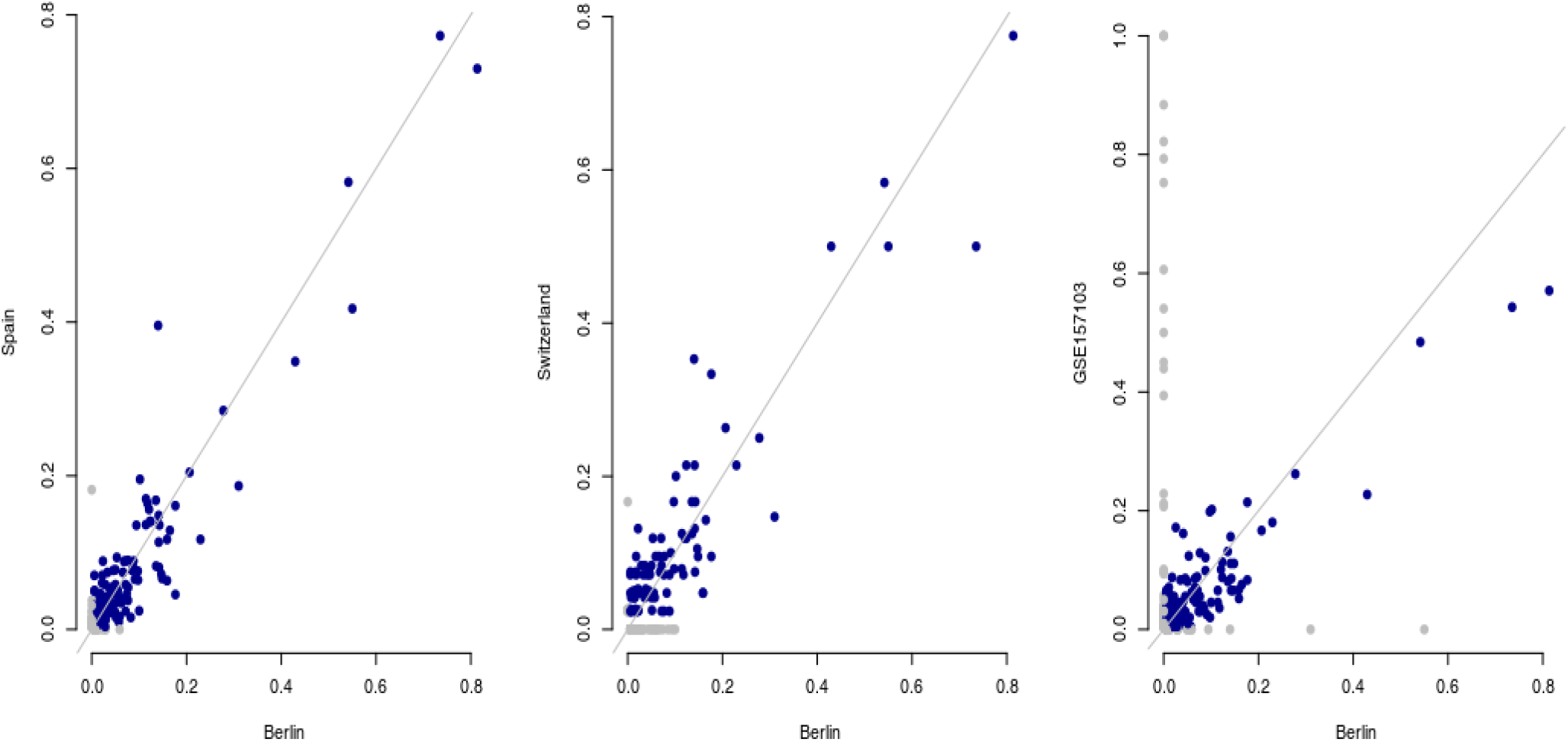

**Supplemental Figure 3.**
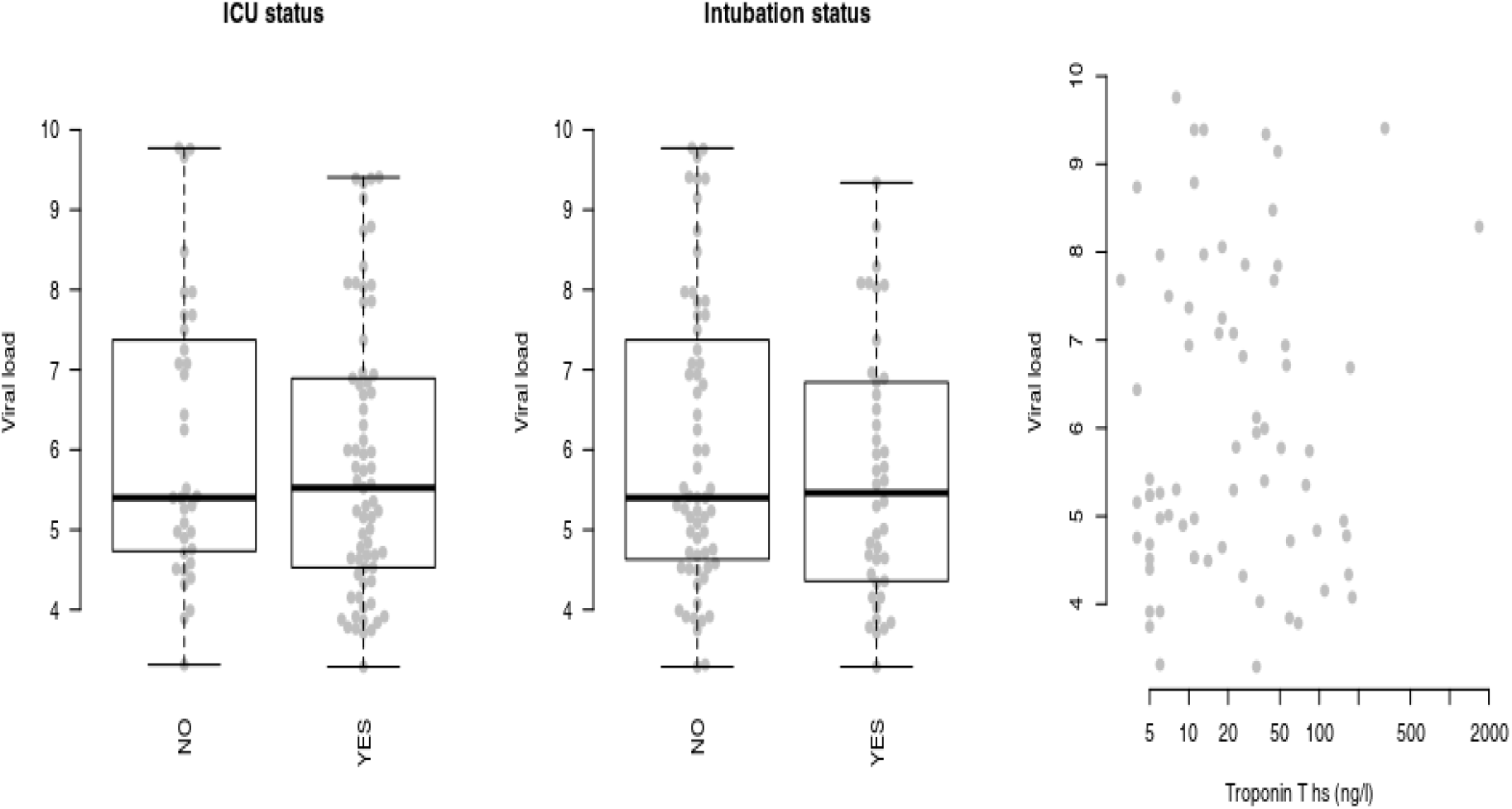
Viral load at first measurement was not significantly associated with ICU / intubation status or troponin T hs levels.

**Supplemental Figure 4.**
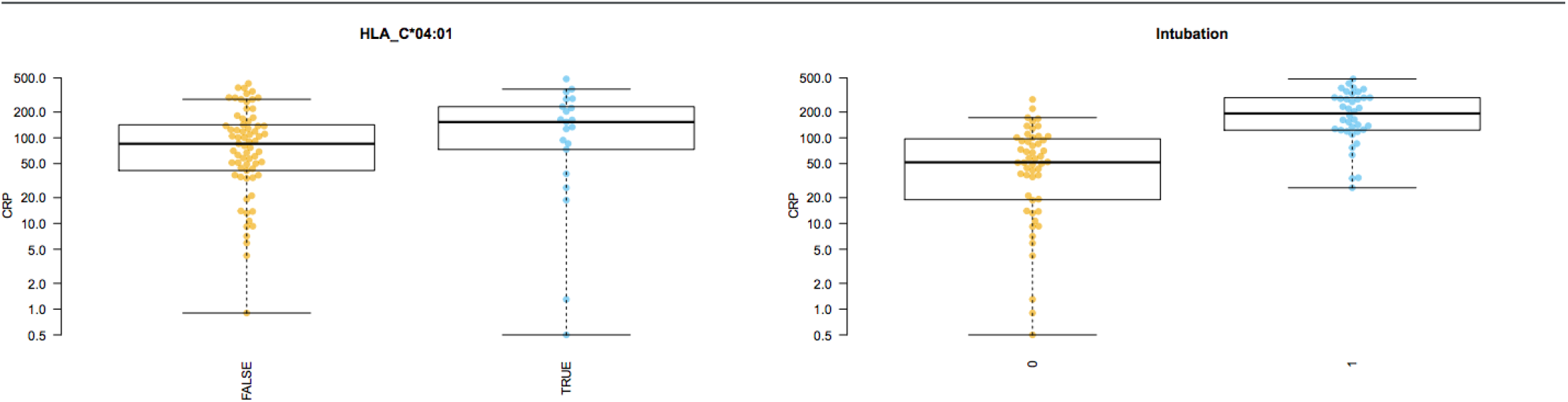
Evaluation for correlation of HLA* C 04:01 status with CRP serum levels. There was a trend of higher CRP levels in patients with HLA* C 04:01. However, these findings were not significant. CRP correlated with the risk of intubation.

## SUPPLEMENTAL TABLES

**Supplemental Table 1.** Significant associations between intubation status and HLA alleles across all data sets. Allele, two-digit allele code; covariate, the covariate for which the association was tested; Frequency: allele frequency; [-/-]: number of individuals not carrying the allele; [-/h,h/h]: number of individuals carrying the allele; Effect size: Cramer’s v d; P-value: p-value from t-test; FDR: p-value corrected for false discovery rate. Only alleles with minimal frequency of 0.05 and present in at least 5 Patients were tested for association.

| *Allele* | *Frequency* | *[-/-]* | *[-/h,h/h]* | *E* | *p.val* | *p.adj* |
| --- | --- | --- | --- | --- | --- | --- |
| *DQA1 03:01* | 0.077 | 273 | 39 | 0.22 | 1e-04 | 0.0069 |
| *HLA_C 04:01* | 0.16 | 251 | 69 | 0.2 | 0.00025 | 0.017 |
| *DQB1 03:02* | 0.11 | 239 | 45 | 0.17 | 0.0038 | 0.26 |
| *DPB1 04:02* | 0.071 | 265 | 36 | 0.16 | 0.0053 | 0.35 |
| *HLA_B 35:01* | 0.052 | 293 | 26 | 0.14 | 0.011 | 0.74 |
| *DRB1 01:01* | 0.055 | 242 | 24 | 0.14 | 0.015 | 0.96 |
| *DQB1 06:02* | 0.079 | 253 | 31 | 0.12 | 0.034 | 1 |

**Supplemental Table 2.** Significant associations in DS1 used to obtain a list of allele candidates. The associations were only tested for alleles with a frequency higher than 5% and present in at least five patients. Parameter, the parameter for which the association was strongest (composite: composite score; intubation status; maximum WHO ordinal scale for clinical improvement score (WHO score); troponine as continuous variable; ICU status). Frequency, frequency of the allele; [-/-]: number of individuals not carrying the allele; [-/h,h/h]: number of individuals carrying the allele; P-value, p-value of the association (t-test for continuous variables, Fisher’s exact test for categorical variables); FDR, p-value corrected for false discovery rate.

| Allele | Parameter | Frequency | [-/-] | [-/h,h/h] | P-value | FDR |
| --- | --- | --- | --- | --- | --- | --- |
| HLA_C 03:03 | composite | 0.070 | 37 | 6 | 0.00023 | 0.0095 |
| HLA_C 04:01 | Intubation | 0.121 | 70 | 21 | 0.00487 | 0.2632 |
| DRB1 11:04 | WHO score | 0.053 | 61 | 5 | 0.01053 | 0.5685 |
| HLA_B 08:01 | WHO score | 0.083 | 70 | 14 | 0.01415 | 0.7499 |
| HLA_C 04:01 | WHO score | 0.121 | 67 | 20 | 0.01976 | 1.0000 |
| DQA1 05:01 | troponin (cont.) | 0.103 | 47 | 11 | 0.02352 | 1.0000 |
| DRB1 01:01 | Intubation | 0.103 | 56 | 12 | 0.02433 | 1.0000 |
| HLA_B 08:01 | troponin (cont.) | 0.086 | 53 | 11 | 0.02433 | 1.0000 |
| DPB1 02:01 | troponin (cont.) | 0.145 | 42 | 13 | 0.03014 | 1.0000 |
| HLA_A 01:01 | Intubation | 0.147 | 67 | 25 | 0.03454 | 1.0000 |
| HLA_B 35:01 | Intubation | 0.074 | 75 | 13 | 0.03821 | 1.0000 |
| DRB1 01:01 | WHO score | 0.098 | 55 | 11 | 0.03918 | 1.0000 |
| HLA_C 01:02 | ICU | 0.082 | 77 | 14 | 0.04173 | 1.0000 |
| HLA_C 04:01 | ICU | 0.121 | 70 | 21 | 0.04387 | 1.0000 |
| HLA_A 01:01 | WHO score | 0.148 | 64 | 24 | 0.04562 | 1.0000 |
| DQB1 03:03 | WHO score | 0.055 | 65 | 8 | 0.04924 | 1.0000 |

**Supplemental Table 3.** Final analysis results. Candidates selected from Data Set 1 were compared to the association results in the validation Data Sets 2 and 3. Only alleles with frequency > 0.05 and present in more than five individuals in all three data sets were considered. P-value was obtained from a randomization testing procedure (replicating the selection and validation process with randomly matched genotypes and clinical data). F1-F3, frequencies in DS1-3; P1-P3, p-values (not adjusted) in DS1-3; P-value, p-value from randomization test; FDR, p-value from randomization test adjusted for multiple testing. Not all alleles were present at required frequency of 0.05 in DS3.

| Allele | F1 | F2 | F3 | P1 | P2 | P3 | P-value | FDR |
| --- | --- | --- | --- | --- | --- | --- | --- | --- |
| HLA_A 01:01 | 0.15 | 0.076 | 0.066 | 0.035 | 0.64 | 0.55 | 0.8 | 1 |
| HLA_B 08:01 | 0.083 | 0.076 | - | 0.014 | 0.62 | - | 0.64 | 1 |
| HLA_C 04:01 | 0.12 | 0.15 | 0.1 | 0.0049 | 0.71 | 0.0012 | 6e-04 | 0.0036 |
| DPB1 02:01 | 0.15 | 0.18 | 0.13 | 0.03 | 0.33 | 0.82 | 0.65 | 1 |
| DQA1 05:01 | 0.1 | 0.16 | - | 0.024 | 0.11 | - | 0.26 | 1 |
| DRB1 11:04 | 0.053 | 0.071 | - | 0.011 | 0.029 | - | 0.042 | 0.21 |

**Supplemental Table 4.** Significant associations between viral load and HLA alleles in data set 1. Allele, two-digit allele code; covariate, the covariate for which the association was tested; Frequency: allele frequency; [-/-]: number of individuals not carrying the allele; [-/h,h/h]: number of individuals carrying the allele; Effect size: Cohen’s d; P-value: p-value from t-test; FDR: p-value corrected for false discovery rate.

| Allele | Covariate | Frequency | [-/-] | [-/h,h/h] | Effect size | P-value | FDR |
| --- | --- | --- | --- | --- | --- | --- | --- |
| HLA_B 15:01 | First measurement | 0.068 | 51 | 8 | 0.64 | 0.0085 | 0.42 |
| HLA_C 03:03 | First measurement | 0.057 | 54 | 7 | 0.79 | 0.0088 | 0.42 |
| DRB4 01:03 | First measurement | 0.66 | 10 | 21 | 1 | 0.02 | 0.92 |
| DRB4 01:01 | First measurement | 0.34 | 20 | 11 | -0.93 | 0.025 | 1 |
| HLA_A 03:01 | Estimated peak viral load | 0.1 | 33 | 7 | 0.57 | 0.034 | 1 |

**Supplemental Table 5.** Associations of HLA alleles and age, gender, weight and BMI. Only alleles which pass the specified thresholds (frequency > 0.05 and occurs in at least five) have been tested for association.

| ***Allele*** | ***Covariate*** | ***Frequency*** | ***Effect size*** | ***P-value*** | ***FDR*** |
| --- | --- | --- | --- | --- | --- |
| *HLA_C 03:04* | age | 0.066 | -0.70 | 0.0047 | 0.25 |
| *DPB1 02:01* | sex | 0.144 | 0.66 | 0.0254 | 1.00 |
| *DQB1 03:02* | weight | 0.128 | -0.77 | 0.0305 | 1.00 |
| *DQB1 06:02* | age | 0.175 | -0.45 | 0.0321 | 1.00 |
| *DRB5 01:01* | age | 0.781 | -1.23 | 0.0474 | 1.00 |
| *DRB5 02:02* | age | 0.188 | 1.23 | 0.0474 | 1.00 |
| *DRB1 15:01* | sex | 0.140 | -0.50 | 0.0481 | 1.00 |

