## Supplementary material for "HLA-C^*^ 04:01 is a Genetic Risk Allele for Severe Course of COVID-19": Cover Letter

### CHARITÉ

#### ChariteCentrum für Herz-, Kreislauf- und Gefäßmedizin

Charité | Campus Benjamin Franklin | 12200 Berlin
Editorial Board
medRXiv

**Klinik für Kardiologie**
**Klinikdirektor**
**Univ.-Prof. Dr. med. Ulf Landmesser**
BIH-Professor für Kardiologie

Hindenburgdamm 30
12203 Berlin

**Kardiologisches Sekretariat**:


Datum: Dec 14, 2020

**Cover Letter**

Dear Editor,

We wish to submit our original article entitled "HLA-C* C04:01 is a Genetic Risk Allele for Severe Course of COVID-19 " to be considered for publication on medRxiv. In our manuscript, we report that patients with the allele HLA-C* 04:01 have twice the risk of intubation, when infected with severe acute respiratory syndrome coronavirus 2 (SARS-CoV-2). This effect was strong (HR 2.1) and highly significant (adjusted p=0.0036).

In March 2020, at the beginning of the coronavirus disease 2019 (COVID-19) pandemic, we established an international European collaboration to test for the association of HLA with clinical course in COVID-19. We performed for the first time full length HLA sequencing of HLA Class I and II alleles using DNA from patients with the full clinical spectrum of COVID-19. We validated our findings successfully in an independent cohort from the United States. To test for biological plausibility, we used clinical and laboratory data, viral load, and performed whole exome sequencing and HLA affinity analysis. Our data suggest a potential predisposition to worsened outcome of patients with HLA-C* 04:01 due to their low binding affinity for SARS-CoV-2 peptides. There was also a trend of higher CRP levels in patients with HLA-C* 04:01, albeit not significant.

Considering the potential impact of HLA-C* 04:01 on clinical course in COVID-19, testing for this allele may be valuable in clinical trials in order to identify which patients benefit most from specific therapies.

On a broader scale, we noted that while the allele frequency of HLA-C* 04:01 is approximately 13%-16% in countries in which our patients were enrolled; it is highest in Mexico (38%, *http://www.allelefrequencies.net*). Interestingly, African Americans are also among the populations with the highest frequency of HLA-C* 04:01 (21%). While socioeconomic and classical risk factors, such as population age and comorbidities, are most likely the strongest predictors for severe clinical outcomes in patients with COVID-19, our data raise the possibility that disease severity may be exacerbated in populations with high HLA-C* 04:01 frequency, contributing to the reported increased frequency of severe outcomes in the African American population. Future studies may eventually determine whether expression of HLA-C* 04:01 is in and of itself sufficient to lead to worsened outcome, or whether it also requires the presence of comorbidities resulting from healthcare disparities, or from other socioeconomic factors.

None of the manuscript’s contents have been published, nor are they under consideration for publication by any other journal other than medRxiv at the current moment. We are planning to submit our manuscript also to a peer-reviewed journal. No portion of this manuscript has been copied from other material in the literature without citation. All authors have participated meaningfully in this work and have seen and approved the final version of the manuscript.

We would like to disclose as potential conflicts of interest that this study was partially funded by Roche Sequencing Solutions, Inc., which also provided material for exome sequencing. We have mentioned involvement by Roche in the acknowledgement section and under disclosures. Data collection and analysis was performed at our university without relevant involvement by Roche. I am an inventor on patents that use RNA for diagnosis of myocarditis and mentioned this under disclosures.

We genuinely hope that our original article is suitable for publication on medRxiv.

Sincerely,

Bettina Heidecker, MD
Head of Heart Failure and Cardiomyopathies Charité
Universitätsmedizin Berlin, Campus Benjamin Franklin
Hindenburgdamm 30
12203 Berlin

Cell: +49 17 360 098 45
